# Decoding Finely-Tuned Gamma Oscillations in Chronic Deep Brain Stimulation for Parkinson’s Disease

**DOI:** 10.1101/2025.05.06.25327071

**Authors:** Marjolein Muller, Eline A.M.Y. Rouleau, Saskia van der Gaag, Rodi Zutt, Carel F. Hoffmann, Niels A. van der Gaag, Thomas A. van Essen, Alfred C. Schouten, M. Fiorella Contarino

## Abstract

**Background:** Finely-tuned gamma (FTG) activity—spontaneous narrowband oscillations within the low-gamma range or entrained to half the stimulation frequency—is traditionally associated with ON medication states and dyskinesia in Parkinson’s disease (PD), representing a potential biomarker for adaptive deep brain stimulation (DBS). However, the conditions in which FTG arises and its determinants remain unclear.

**Methods:** Local field potentials recorded in the subthalamic nucleus (STN) in PD patients with SenSight^TM^ leads and Percept^TM^ neurostimulator were retrospectively analyzed. The analysis was guided by a comprehensive list of clinically relevant questions.

**Results:** Among 67 patients (134 STN), spontaneous FTG was observed in 11% of STNs, and entrained FTG—including both 1:2 and the novel 1:1 entrainment— in 33%. FTG types never appeared simultaneously and were mostly ON-related but independent of dyskinesia. Beta-activity was inversely related with 1:2 entrainment during stimulation state transitions. Notably, as previously hypothesized, 1:2 entrainment diminished with higher stimulation amplitudes and was inversely related to 1:1 entrainment.

**Conclusions:** This study provides clinical evidence of 1:1 entrainment. FTG expression – spontaneous, 1:2, or 1:1 entrained – is modulated by DBS and clinical state, but can occur independent of dyskinesia. We confirm that FTG is prokinetic and is inversely related with akinetic beta activity.

## 1. Introduction

Deep brain stimulation (DBS) of the subthalamic nucleus (STN) has become an established therapy for motor symptoms in Parkinson’s disease (PD), especially for patients refractory to pharmacological treatments [1]. Although PD patients benefit significantly from DBS, the programming and optimization process can be burdensome, and its subjectivity can introduce and explain variability in patient outcomes [2]. To address this challenge, novel neurostimulation systems allow the chronic recording of local field potentials (LFP) directly from the implanted neurostimulators [3]. LFP represent the integrated activity of synaptic potentials and provide insight into the functioning of basal ganglia circuits [4, 5]. The possibility of recording LFP from the implanted leads presents the opportunity to further refine the initial programming and the ongoing adjustments of DBS, offering a more objective, data-driven approach [6–9]. As potential neurophysiological biomarkers (physiomarkers), LFP could guide more precise and individualized DBS therapy adjustments and form the basis for adaptive DBS (aDBS). [10, 11].

Research on LFP has traditionally focused on the beta frequency band (13-35Hz), which correlates with bradykinesia and rigidity in PD [12, 13]. Beta oscillations are modulated by treatments such as levodopa and STN-DBS, representing a potential physiomarker for DBS programming [10, 11]. However, establishing fully automated and reliable DBS protocols based merely on beta frequency activity remains challenging [14–16]. Consequently, recent interest has turned toward the prokinetic gamma frequency band. Specifically, finely-tuned gamma (FTG) activity, defined as narrowband (i.e. ≤10Hz) activity within the low-gamma range, has been shown to correlate with medication- or stimulation-induced ON state and dyskinesia in PD patients [17–21].

The use of FTG provides a possible avenue for further improving DBS programming and aDBS stimulation protocols, independently or in combination with beta-activity [17, 22, 23]. FTG has been reported to occur both as spontaneous activity (spontaneous FTG - sFTG) and as an entrained response to stimulation (entrained FTG - eFTG) [21, 24]. This entrainment is typically detected at half the stimulation frequency (1:2 entrainment), creating predictable patterns associated with ON periods during DBS therapy [17, 25, 26]. Nevertheless, the mechanisms underlying FTG remain poorly understood, in particular, the conditions under which it emerges and becomes entrained by stimulation (i.e. medication state, stimulation parameters, and the presence of specific symptoms such as dyskinesia) are not fully clarified due to limited and contradicting research on this topic [17, 23, 24, 26].

Recently, the ‘Arnold’s tongue’ framework has been proposed to describe the mechanisms underlying the entrainment of FTG. This framework describes how an oscillatory system can become entrained to an external periodic stimulus, depending on the strength and frequency of the input. In neural systems, it illustrates the range of stimulation parameters over which brain rhythms lock to stimulation at integer or rational frequency ratios (e.g., a frequency at 1:1, 1:2, 1:4, or 3:4 of the stimulation frequency). This concept may help explain how DBS can entrain neural oscillations like FTG. However, this mechanism has not yet been fully validated in PD patients, as an important element of the framework includes the transition of 1:2 entrainment to other frequencies, such as the stimulation frequency (1:1 entrainment).

This has not yet been established in clinical recordings [25, 26] due to the prominent presence of a stimulation artifact. Furthermore, since the eFTG described so far corresponds to a subharmonic of the stimulation frequency (1:2), the observed entrainment may also reflect a stimulation artifact instead of a physiological response [16, 27]. This might be particularly true for the potential 1:1 entrainment.

In this study we aim to further clarify the characteristics of FTG in PD patients with STN-DBS, with a particular focus on its prevalence and relationship with patient characteristics and stimulation parameters as observed in daily practice. Furthermore, we explore the relationship between 1:2 and 1:1 entrainment as well as the relation of FTG with established effects of stimulation on the beta frequency band. By reporting these daily practice observations, we wish to strengthen and challenge some of the current assumptions regarding the clinical associations and origin of FTG in PD.

## 2. Materials and Methods

### 2.1 Study design

This retrospective observational cohort study included PD patients who underwent DBS surgery and LFP recordings between January 1^st^, 2020 and June 1^st^, 2023. The Medical Ethical Committee Leiden-Den Haag-Delft waived formal review (reference N23.007). In accordance with the standard opt-out procedure, all patients were informed and given the opportunity to object to the use of their data for research purposes.

Eligible patients had bilaterally implanted directional leads (SenSight™ B33005, 1-3-3-1 design with 0.5mm spacing between contacts) in the STN and a sensing-enabled neurostimulator (Percept^TM^ PC, Medtronic, Minneapolis). Inclusion required at least one BrainSense^TM^ recording within the study period.

All available recordings from included patients, acquired during routine clinical care within the study period, were collected and analyzed. A structured list of clinically relevant questions was developed to guide the analysis and highlight key aspects of FTG. These questions were addressed using the available retrospective data:

### Associations between FTG, clinical state and dyskinesia

a. Does sFTG / eFTG only occur in association with dyskinesia?
b. In which clinical state does sFTG / eFTG occur? (ON/OFF-medication, ON/OFF-stimulation, or during the stun effect period (i.e. <1 month post-operative period in which (subclinical) temporary improvement of motor symptoms due to a surgically created microlesion can occur).
c. If sFTG / eFTG also occurs in the OFF state (OFF-medication / no stun effect expected ≥1 month post-operative), is this associated with the presence of tremor?

### Characteristics of spontaneous FTG and entrained FTG

a. At what frequency band(s) does sFTG / eFTG occur?
b. What is the bandwidth of this sFTG / eFTG activity?
c. Is the power of sFTG / eFTG (with the same stimulation amplitude) stable or does it fluctuate over time?
d. If it fluctuates, how long does sFTG / eFTG last in a single recording?
e. Is eFTG modulated by different stimulation amplitudes or stimulation frequencies?
f. If present, is sFTG / eFTG always visible in all contact pairs in one hemisphere or is it location-specific?
g. Is there an influence of unilateral stimulation in the contralateral non-stimulated hemisphere?

### Relation between spontaneous FTG, entrained FTG and beta

a. Is eFTG only observed in recordings and channels where the sFTG was also present?
b. If eFTG follows sFTG, does the eFTG start at the frequency of sFTG?
c. Is sFTG / eFTG in ON medication state only observed in recordings and channels where there was a beta in OFF medication state, and do they correspond in relative power?
d. If sFTG / eFTG follows beta, does it start at the same time when the beta stops, and is there a relationship in the variation in amplitude between beta and sFTG or eFTG?

### 2.2 Local field potential and clinical data collection

Contact-level bipolar LFP recordings were obtained for the virtual rings (i.e. rings including three directional segments) and conventional rings of the SenSight^TM^ lead. These recordings were conducted according to the standard of care, with the patient seated in either the OFF or ON medication state, depending on the clinical requirements. Recordings took place at various time points after surgery, including during the standardized monopolar review approximately 10-days post-operative, as well as during other routine follow-up visits starting from the first day to several years after surgery.

BrainSense^TM^ recordings were visualized using customized Matlab scripts (version R2022b, MathWorks®). The visualized recordings were screened to identify the presence or absence of FTG or broadband gamma. FTG was defined as visible narrowband (i.e. ≤10Hz wide) activity within the low-gamma range (40Hz-125Hz, as higher frequencies could not be recorded with the used system). Whereas, broadband gamma was defined as visible broadband (i.e. >10Hz wide) activity within the same frequency range. Both FTG and broadband gamma could be visually distinguished from background activity.

A further distinction was made between “Spontaneous FTG”, defined as FTG occurring at frequencies unrelated to the stimulation frequency, and “Entrained FTG”, characterized by activity centered at precisely half the stimulation frequency (1:2 entrainment) or at the stimulation frequency itself (1:1 entrainment). Here, activity at the stimulation frequency was classified as 1:1 entrainment only when power variations at this frequency did not correlate with changes in stimulation amplitude. If activity at the stimulation frequency varied solely in accordance with the stimulation amplitude, the activity was considered a stimulation artifact.

When FTG was detected, multiple features were documented, including stimulation parameters (frequency, amplitude, pulse width, active contact-level), the recording channel of occurrence, FTG frequency and amplitude and any fluctuations thereof. Relevant clinical characteristics at the time of each recording were extracted from electronic patient records, including medication state, symptom presence (tremor, rigidity, bradykinesia, and dyskinesia), and time elapsed since lead implantation.

### 2.3 Local field potential pre-processing and analysis

Time-domain data (sampling frequency 250Hz) from the BrainSense^TM^ Survey, Setup, Indefinite Streaming, and Streaming recordings [28] were transformed to the frequency domain for visualization using power spectral density plots and spectrograms. These analyses employed methods form the existing “PerceptToolbox” in Matlab (v. 2023b) [16]. Spectral density plots were computed using Welch’s method over the frequency range of 1–125Hz.

The signals were segmented into 5-second Hamming windows with 60% overlap, providing a frequency resolution of 0.1Hz. For time-frequency analysis, spectrograms were generated using the short-time Fourier transform with 0.5-second Hamming windows and 50% overlap, also covering the 1–125Hz frequency range. BrainSense^TM^ Events, which are stored exclusively in the frequency domain, were also visualized using a modified version of the existing scripts.

For recordings permitting changes in stimulation amplitude during the recording session (BrainSense^TM^ Streaming recordings), z-scores were determined and visualized for the beta-band (15-30Hz), sFTG, 1:2 eFTG, and, if possible, for 1:1 entrainment or stimulation frequency artifacts (activity at 120-125Hz). Normalization procedures involved comparing beta-band activity to the broader 5-45Hz range; spontaneous and eFTG activity was normalized against part of the low-gamma-band (40-100Hz); and, in the case of stimulation at 125Hz, activity at the stimulation frequency was compared to activity within the 110- 120Hz band.

## 3. Results

### 3.1 Study population

In 67 patients (134 STN: 3389 n=12, SenSight^TM^ B33005 n= 122) at least 1 recording was performed within the study period. During these recordings, stimulation was toggled OFF (all 134 STN), or ON with stimulation at 125Hz (118 STN), 130Hz (4 STN) or 180Hz (20 STN), where 8 of the STN were recorded both during 125Hz and 180Hz stimulation. **Figure 1** provides a summary of the amount of STN showing different forms of gamma activity in varying stimulation and recording conditions.

**Figure 1.**
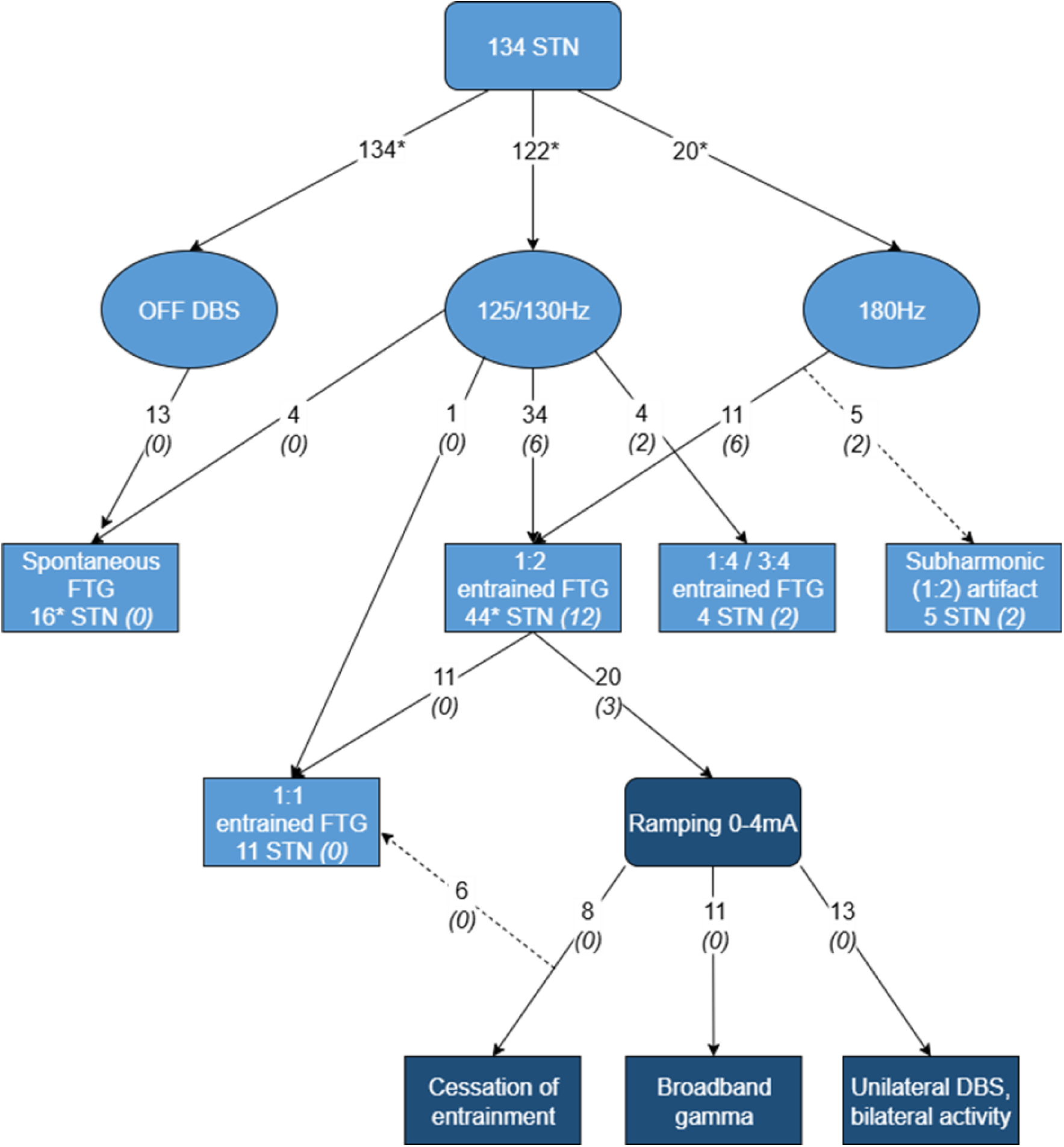
Flowchart providing an overview of the amount of STN showing different gamma activities across all the available BrainSense^TM^ recordings with relation to the stimulation state. Recordings were available for 134 STN in total. Of these STN all had recordings OFF DBS, and most also ON DBS, either with 125/130Hz or 180Hz stimulation. For these three DBS states (OFF; ON-125/130Hz; ON-180Hz) the flowchart provides the amount of STN showing spontaneous finely-tuned gamma (FTG) (i.e. narrowband activity between 40-125Hz at a semi-random frequency); 1:1 entrainment (i.e. activity at the stimulation frequency modulated independent from the stimulation amplitude); 1:2 entrainment (i.e. FTG activity entrained to ½ the stimulation frequency); 1:4 / 3:4 entrainment (i.e. FTG activity entrained to ¼ and/or ¾ the stimulation frequency); or a subharmonic artifact (i.e. activity at exactly ½ the stimulation frequency persisting unchanged with stimulation amplitude of 0mA). For some of the cases showing 1:2 entrainment, stimulation was ramped from 0mA to 4mA and back to 0mA. In some of these ramping cases one of the following was observed: cessation of 1:2 entrainment during the ramping up or anyway prior to a decrease in stimulation amplitude; presence of broadband gamma activity (i.e. broadband activity between 40-100Hz); or presence of bilateral activity at the stimulation frequency during unilateral DBS. *(#)* Count of STN recorded in the OFF medication state without expected stun effect (≥1 month post-operative). * Several STN were recorded in multiple stimulation states.

In 31 of the patients (51 STN, 38%), FTG was identified in at least one recording. sFTG was visible in 10 patients (6 bilateral and 4 unilateral), of which 7 patients (2 bilateral, 5 unilateral) also showed eFTG. Traditional 1:2 eFTG occurred in 28 patients (16 bilateral and 12 unilateral). Notably, 11 of these STN also exhibited the previously clinically unverified 1:1 entrainment. The clinical characteristics of patients with and without FTG are shown in **Table 1**.

**Table 1.** Clinical characteristics of patients with and without spontaneous and/or entrained finely-tuned gamma activity.

|  | Spontaneous FTG<br>(N = 10 patients)<sup>3</sup><br>Median (range) | Entrained FTG<br>(N = 28 patients)<sup>3</sup><br>Median (range) | Bilateral no FTG<br>(N = 36)<br>Median (range) |
| --- | --- | --- | --- |
| Age at lead implant, years | 63 (50-69) | 66 (41-79) | 61 (49-76) |
| Sex (male), N (%) | 6 (60%) | 21 (75%) | 18 (50%) |
| Lead type, N (%):<br>- 3389<br>- SenSight™ | 0 (0%)<br>10 (100%) | 1 (4%)<br>27 (96%) | 5 (14%)<br>31 (86%) |
| Percept PC, N (%):<br>- Primary implant<br>- Replacement | 10 (100%)<br>0 (0%) | 28 (100%)<br>0 (0%) | 33 (92%)<br>3 (8%) |
| Disease duration at lead implant, years | 9 (4-20) | 7 (2-20) | 9 (3-21) |
|  | Spontaneous FTG<br>(n = 16 STN)<sup>4</sup> | Entrained FTG<br>(n = 44 STN)<sup>4</sup> | No FTG<br>(n = 1+10+72 STN)<sup>5</sup> |
| Main Symptom<sup>1</sup>, n (%): |  |  |  |
| Tremor only | 1 (6%) | 4 (9%) | 0+3+3 (7%) |
| Tremor combined<sup>2</sup> | 5 (31%) | 18 (41%) | 0+2+18 (24%) |
| Bradykinesia/rigidity only | 7 (44%) | 18 (41%) | 1+3+44 (58%) |
| Stun effect (no symptom) | 3 (19%) | 4 (9%) | 0+2+7 (11%) |
| Time since implant at recording, days | 75 (0-549) | 56 (0-1220) | 59 (0-1220) |

<sup>1</sup> Main symptom per hemisphere based on MDS UPDRS-III scored in OFF-medication and OFF-stimulation state at 8-10 days post-operative (during monopolar review).
<sup>2</sup> Tremor with similar MDS UPDRS-III score as bradykinesia and/or rigidity.
<sup>3</sup> Patients exhibiting both spontaneous and entrained FTG were included in both subgroups (N = 7).
<sup>4</sup> STN exhibiting both spontaneous and entrained FTG were included in both subgroups (n = 9).
<sup>5</sup> STN without FTG divided with respect to the original patient group (STN from patients originally classified as patients with spontaneous FTG + STN from patients originally classified as patients with entrained FTG + STN from patients originally classified as patients with bilaterally absent FTG)
*Abbreviations:* FTG: finely-tuned gamma; N: number of patients; MPR: monopolar review; STN: subthalamic nucleus; n = number of STN.

### 3.2 Associations between FTG, clinical state and dyskinesia

#### 3.2.1 Spontaneous FTG

All cases of sFTG were either recorded ON-medication (n = 11 STN) or during the stun effect period (<1 month post-operative, n = 5 STN). sFTG occurred mostly in the OFF-stimulation state. However, in 4 out of 16 STN (25%) sFTG was present ON-stimulation with a maximum stimulation amplitude between 1.2 and 2.6mA.

Although sFTG was often associated with dyskinesia (80%), two patients were without dyskinesia during sFTG recording (ON-medication in one patient, and OFF-medication but with a clinical stun effect in another patient). Additionally, not all patients experiencing dyskinesia displayed sFTG (**Figure 2**).

**Figure 2.**
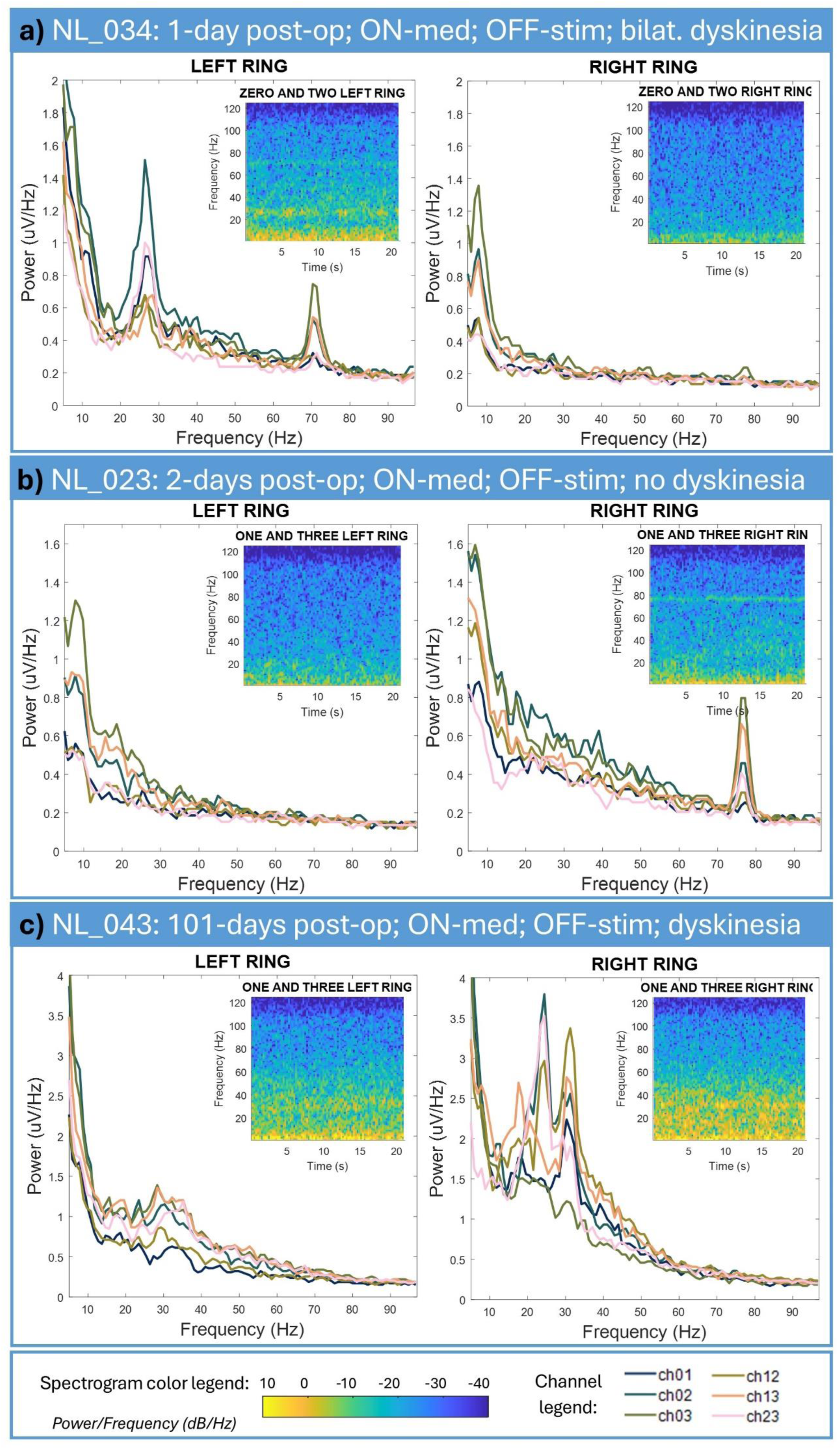
Spontaneous finely-tuned gamma (FTG). Power spectral density plots for local field potential recordings performed using BrainSense^TM^ Survey in both hemispheres of three individuals during in-clinic visits. All recordings were performed in the ON-medication and OFF-stimulation states either with or without the patient experiencing dyskinesia. The spectrogram for the channel surrounding the currently used stimulation contact is also shown. NL_034 **(a)** was recorded 1 day post- operative while experiencing dyskinesia in both arms. The recording showed high beta and FTG activity in the left hemisphere, whereas the right hemisphere showed no beta-activity (stun effect) and only limited spontaneous FTG (sFTG) activity. NL_023 **(b)** was recorded 2 days post-operative and showed no distinct beta-activity in either hemisphere but strong sFTG activity in the right hemisphere and limited sFTG activity in the left hemisphere without experiencing dyskinesia. Finally, NL_043 **(c)** was recorded 101 days post- operative. Interestingly, the recording showed low amounts of beta-activity in the left hemisphere, and remarkably high amounts of beta-activity in the right hemisphere, but no sFTG activity whilst experiencing dyskinesia.

#### 3.2.2 Entrained FTG

eFTG (1:2 entrainment or 1:1 entrainment) was mostly observed in the ON-medication state; however, 12 STN (27%) with eFTG were recorded in the OFF-medication state and outside the expected stun effect period (≥1 month post-operative). Among these 12 STN, 6 were stimulated at 180Hz and all exhibited tremor, but none had tremor as the only primary symptom. The only case in which tremor was the only primary symptom occurred in an STN stimulated at 125Hz. In 7 of the 12 STN with eFTG OFF-medication, tremor was present but accompanied by bradykinesia and rigidity of similar severity. In the remaining 4 STN, no tremor was observed.

All patients stimulated at 180Hz had tremor. In contrast, among those stimulated at 125Hz, 1:2 entrainment was not specifically associated with tremor as a primary symptom.

Specifically, 29% of STN with tremor showed 1:2 entrainment at 125Hz, a proportion comparable to the overall rate of 1:2 entrainment observed in the population receiving 125/130Hz stimulation.

eFTG did not show a clear relationship with dyskinesia; 14 patients (50%) with 1:2 entrainment did not experience dyskinesia at the time of the recordings. Of these patients 6 also showed 1:1 entrainment, representing 67% of the population with 1:1 entrainment.

### 3.3 Characteristics of spontaneous FTG and entrained FTG

#### 3.3.1 Spontaneous FTG

sFTG activity was observed within the 63Hz to 110Hz frequency range, with a median frequency spanning from 76Hz to 83Hz. The bandwidth of this activity ranged between 5Hz and 9Hz. The intensity and frequency-location of the sFTG signal slightly varied over time within a single recording (**Figure 2-A/B and Figure 6**) as well as between recording sessions. If sFTG was present in the OFF-stimulation state, it lasted throughout the entire recording, although the occurrence and power varied between contact pairs (**Figure 2**).

#### 3.3.2 Entrained FTG

##### 1:2 entrainment

Within the studied recordings, 1:2 entrainment with a bandwidth of approximately 5Hz was observed in 44 STN. In three additional patients (5 STN) stimulated at 180Hz (5/20, 25%), activity at half the stimulation frequency was present, however, in these cases the activity did not disappear with stimulation at 0mA and was thus considered artifactual (**Figure 3-B**). The persistence of artifact activity can be explained by a characteristic of the neurostimulator which includes hardware switching of the stimulation even at 0mA if the stimulator is ON. This issue would not present with the stimulation completely switched OFF [29], but unfortunately, no recordings were available for these patients in the Streaming mode with stimulation toggled OFF. In all patients stimulated with 125/130Hz 1:2 entrainment did not persist at 0mA.

**Figure 3.**
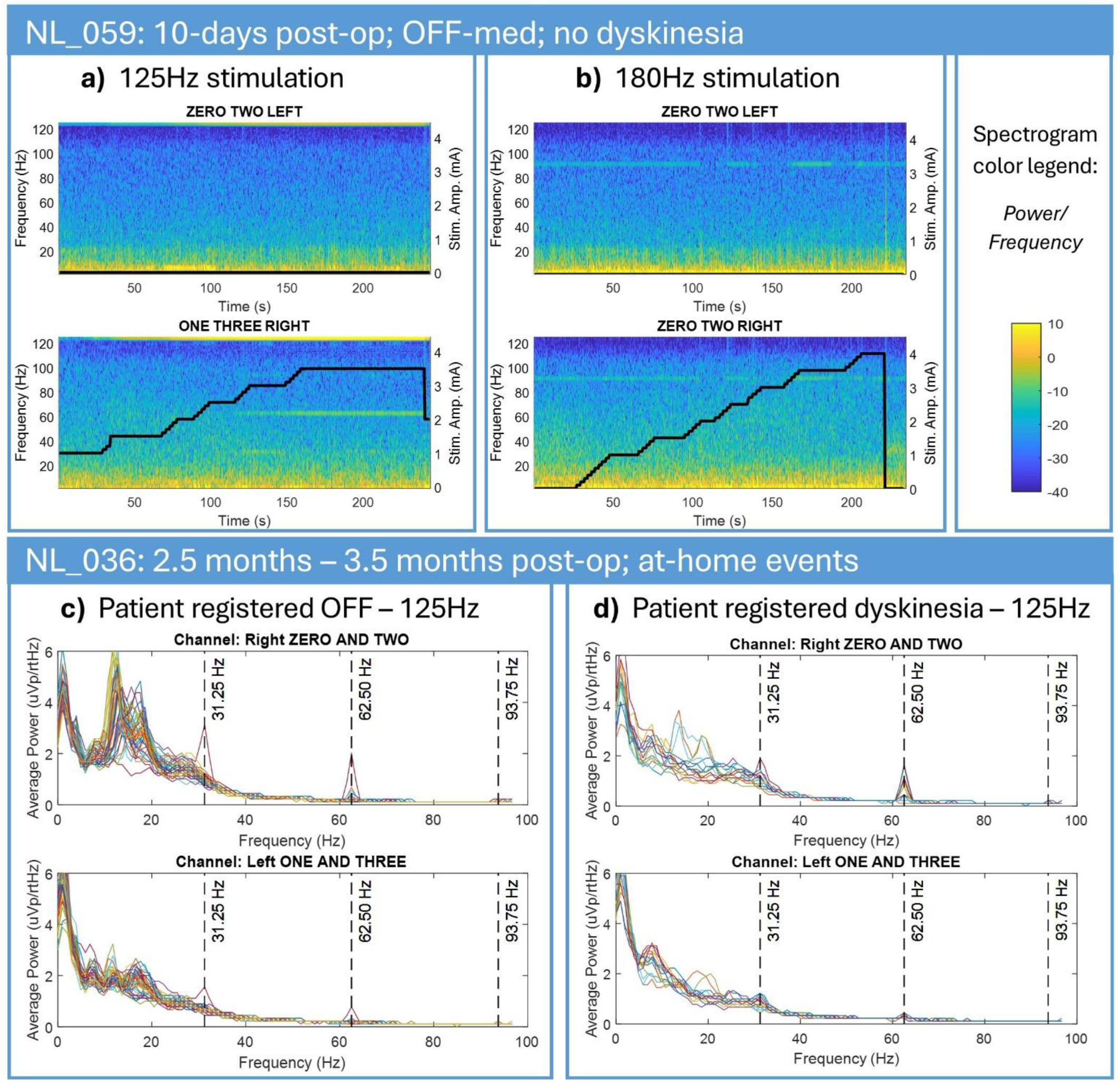
(**a and b**) Local field potential recordings performed during a single in-clinic visit (NL_059) 10 days after surgery in the OFF-medication state. **(a)** the LFP power across 5 to 125Hz during 125Hz stimulation in the channel 1-3 of the right hemisphere starting at 1mA and slowly ramped to 3.5mA and back to 2mA. In the right hemisphere clear activity at half the stimulation frequency (1:2 entrainment) emerges when stimulation reaches 2.6mA. Next to this, activity at 1/4^th^ and possibly 3/4^th^ of the stimulation frequency is apparent. These subharmonics seemingly stop when stimulation is reduced to 2mA. In addition activity at the stimulation frequency is present and seems modulated by the stimulation amplitude (stimulation artifact). Stimulation in the left hemisphere is consistently kept at 0mA, nonetheless activity around the stimulation frequency (125Hz) is present and modulated similarly to the contralateral side, suggesting relation with stimulation changes in the right hemisphere. **(b)** Power across 5 to 125Hz during 180Hz stimulation in the right hemisphere for channel 0-2 at 0mA which is slowly ramped to 4mA before switching back to 0mA. Stimulation in the left hemisphere is consistently kept at 0mA. In both hemispheres a clear activity with mostly constant amplitude at half the stimulation frequency is visible throughout the entire recording independent from the stimulation amplitude and can be interpreted as subharmonic stimulation artifact. At certain timepoints vertical artifacts (activity throughout all frequencies due to impulse noise) are visible, possibly causing the disruptions in the activity at half the stimulation frequency. Activity at the stimulation frequency could not be interpreted here, as 180Hz is not within the recording range. **(c and d)** Activity at 1/2^nd^, 1/4^th^ and 3/4^th^ of the stimulation frequency was also captured in some of the at-home registered OFF events **(c)** and Dyskinesia events **(d)** in another patient (NL_036) stimulated at 125Hz between 2.5 and 3.5 months post-operative.

STN stimulated at 180Hz exhibited 1:2 entrainment more frequently (in 11/20, 55%, of which 1 STN also stimulated at 125Hz) than STN stimulated at 125Hz (33/118, 28%, of which 1 STN also stimulated at 180Hz) or 130Hz (1/4, 25%). The required stimulation amplitude to produce 1:2 eFTG varied across patients, hemispheres, and stimulation frequency. The median stimulation amplitude required for 1:2 entrainment was lower with stimulation at 180Hz (1.3mA, IQR: 0.8mA) compared to 125/130Hz stimulation (2.6mA, IQR: 1.1mA).

Additionally, in one patient, 1:2 entrainment only occurred in a single hemisphere during 125Hz stimulation and in both hemispheres with 180Hz stimulation.

In three patients (4 STN), stimulation at 125Hz resulted in 1:2 entrainment, as well as activity at 1:4 and 3:4 of the stimulation frequency (**Figure 3-A/C/D**).

##### 1:1 entrainment

In case of a stimulation artifact as defined above, the power around the stimulation frequency (here 125Hz) correlates with the changes in the stimulation amplitude (**Figure 3**). However, 12 STN (27%) in our population showed a variation in power around 125Hz which did not correlate with changes in the stimulation amplitude. We consider this a 1:1 entrainment effect (**Figure 4 and Figure 5-A2/C1/C2**). In these cases, an increase in 1:2 entrainment power often corresponded with a decrease in 1:1 entrainment power, and vice versa, irrespective of stimulation amplitude, implicating an inverse relation. In only 1 out of 12 STN, no 1:2 entrainment occurred within the same recording. In cases without variation in power around 125Hz, the presence of 1:1 entrainment could not be excluded due to the potentially larger influence of a stimulation artifact.

**Figure 4.**
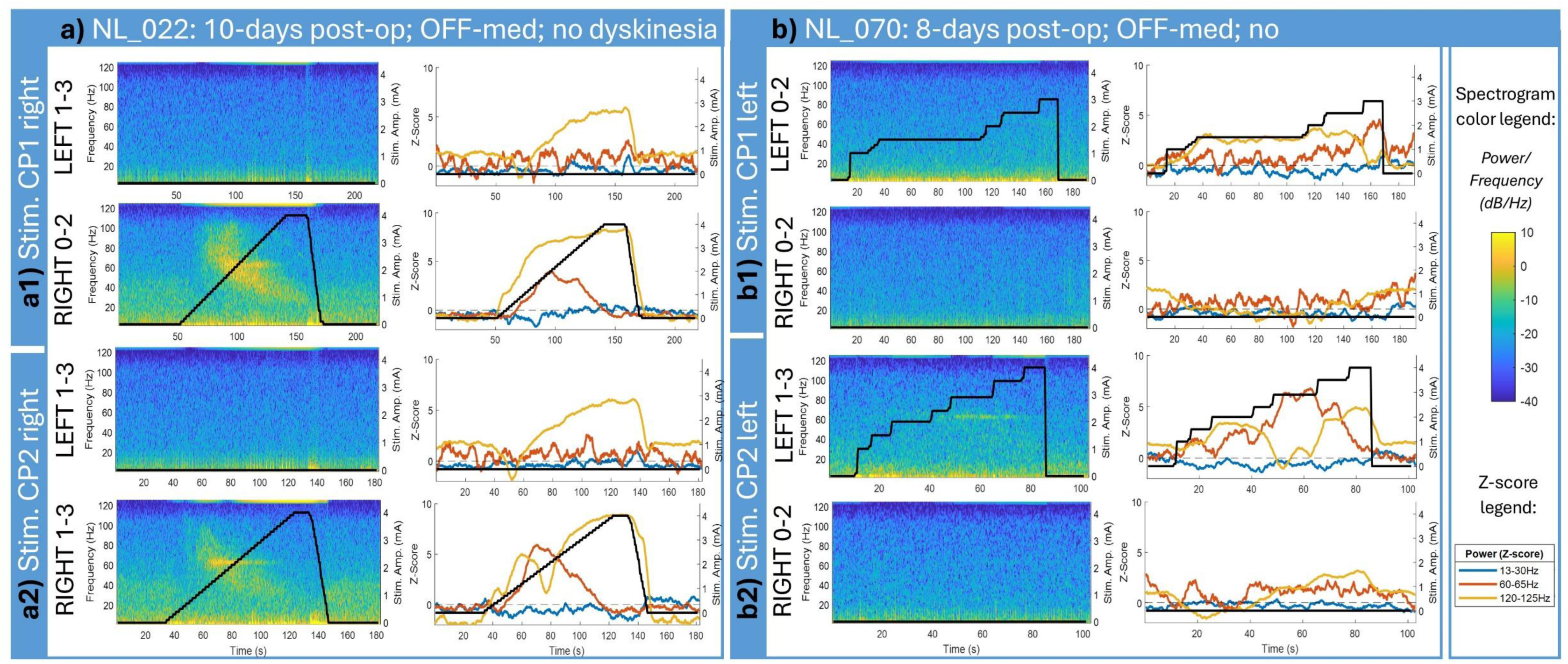
Local field potential recordings from patients NL_022 **(a)** and NL_070 **(b)** during in-clinic visits using the BrainSense Streaming function at 10 and 8 days post-op, respectively, in OFF-medication and ON-stimulation (125Hz) states. Neither patient exhibited dyskinesias during recordings. The left column displays spectrograms, and the right column shows z-scores for beta frequencies (13–30Hz), entrained FTG (eFTG) (60–65Hz), and stimulation amplitude (120–125Hz). For patient NL_022, stimulation was applied in the right hemisphere via contact points 1 **(a1)** and 2 **(a2)**, ramped from 0 to 4mA and back. The left hemisphere was unstimulated. Stimulation at these contacts led to a decrease in beta z-scores, coinciding with increased 1:2 eFTG z-scores. Notably, stimulation at contact point 2 showed an increase in 1:2 eFTG with a corresponding decrease in activity at the stimulation frequency, independent of stimulation amplitude, indicative of a 1:1 entrainment effect. This 1:1 entrainment inversely correlated with the 1:2 entrainment, suggesting competitive interaction. Conversely, stimulation at contact point 1 increased power at the stimulation frequency alongside amplitude increases, considered an artifact rather than 1:1 entrainment. In patient NL_070, stimulation was applied in the left hemisphere at contact points 1 **(b1)** and 2 **(b2)**, ramped from 0 to 3mA and 4mA, respectively, with no stimulation in the right hemisphere. Both contacts elicited 1:2 entrainment and beta-power decreases correlated with stimulation amplitude. A competitive interaction between 1:2 entrainment and activity at the stimulation frequency (1:1 entrainment) was again observed. In all cases, activity at the stimulation frequency also appeared in the contralateral non-stimulated hemisphere, showing artifact patterns similar to the stimulated side but at lower power and without modulation by 1:1 entrainment, even when present in the stimulated hemisphere.

**Figure 5.**
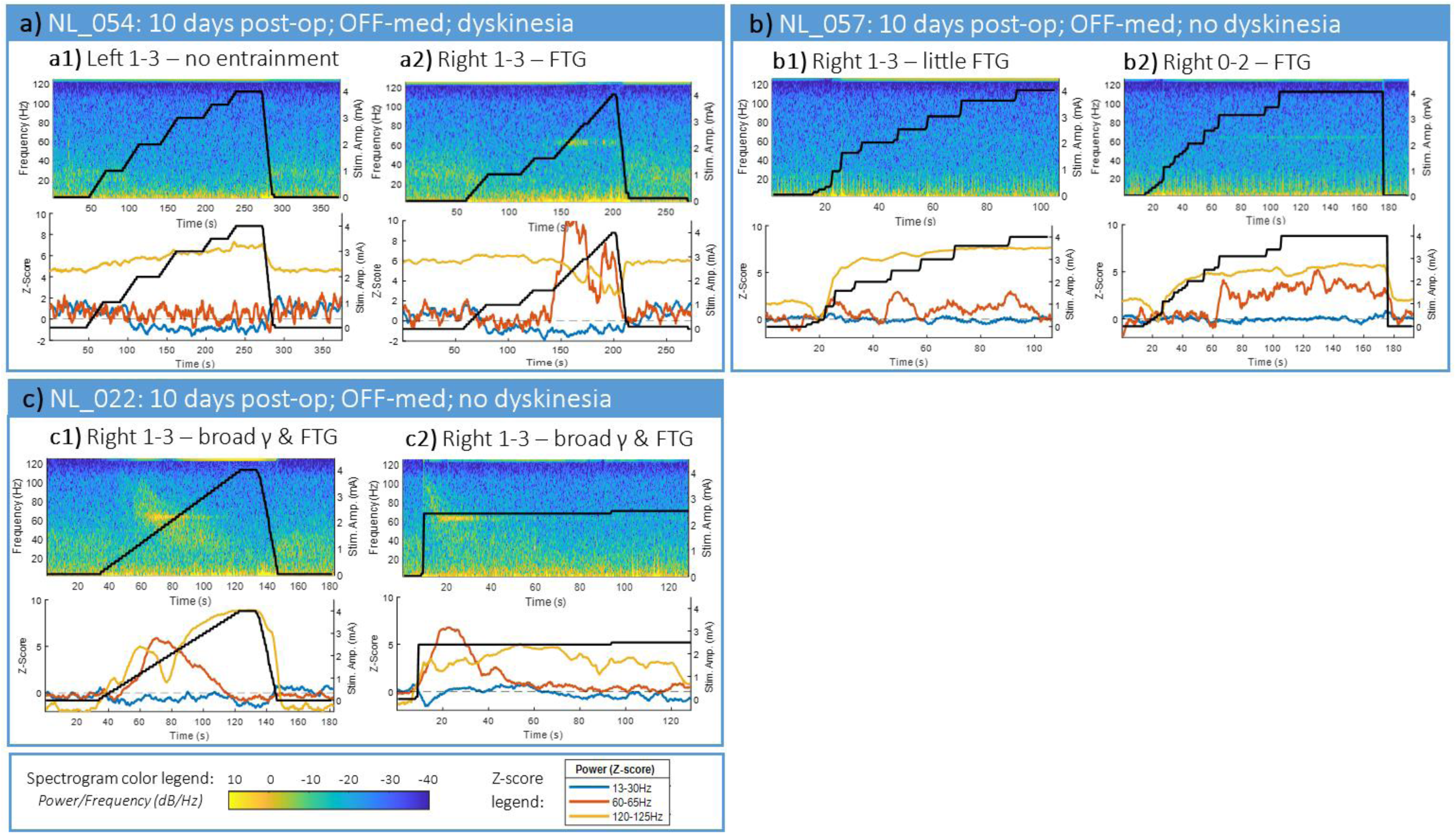
Spectrogram for local field potential recordings performed in three individuals during in-clinic visits using the BrainSense^TM^ Streaming function 10 days after surgery in the OFF-medication and ON-stimulation (125Hz) states. **(a1, b1)** Show recordings performed across a single channel in a single hemisphere for two individuals (NL_054 and NL_057), in A1 no 1:2 entrained FTG (eFTG) is visible, for B1 little 1:2 eFTG is visible. In both cases activity at the stimulation frequency is likely at least partially artifactual, as it is clearly related to the stimulation amplitude. **(a2, b2)** Show recordings from the same patients at the same moment in time, but for a different hemisphere in NL_054 or a different channel in NL_057: here both recordings show 1:2 eFTG across a narrow band between 60-65Hz, the narrow band shows intensity changes over time which, for A2, clearly correspond with changes in activity at the stimulation frequency (1:1) and not with the stimulation amplitude. These changes can this be considered as 1:1 entrainment. The power of this 1:1 entrainment shows clear inverse relation with the power of 1:2 entrainment. Finally, **(c1, c2)** show recordings performed in a different patient (NL_022) for a short (left) and more extended stimulation period (right) in the same channel and hemisphere. Here in both cases, 1:2 eFTG occurs as a narrow band between 60-65Hz but gamma activity is also evident throughout a broad range between 40-110Hz (broadband gamma activity). The intensity of the 1:2 eFTG diminishes before the stimulation amplitude is reduced and shows a clear inverse relation with the activity at the stimulation frequency. This activity at the stimulation frequency varies in power irrespective of the stimulation amplitude, and is thus again considered a 1:1 entrainment effect.

The intensity of the entrained 1:2 FTG power fluctuates over time within a single recording, as well as between recording sessions, partially independent of stimulation amplitude, but occasionally related to 1:1 entrainment power (**Figure 4-A2/B1/B2 and Figure 5-A2/C1**). BrainSense™ Streaming recordings with stimulation ramping from 0mA to 4mA and back to 0mA were available for 20 STN with 1:2 entrainment. In 8 of these STN recordings (40%), the 1:2 entrainment diminished before stimulation was reduced (**Figure 5**). Across 6 of these STN, cessation of 1:2 entrainment led to 1:1 entrainment (in 9 /12 recording channels). The median stimulation amplitude at which 1:2 entrainment disappeared or transitioned was 3.0mA (IQR: 0.6mA).

In most cases, 1:2 entrainment was observed across all contact pairs within a single STN in the same session. However, in 5 out of 44 STN (11%), 1:2 entrainment only occurred in some of the contact pairs. In all 10 patients (13 STN), for which recordings from both hemispheres were available during 125Hz stimulation ramping from 0mA to 4mA, unilateral stimulation led to bilateral activity at the stimulation frequency (**Figures 3 and 4**). Activity in the non-stimulated hemisphere did not vary independently of the stimulation amplitude, despite the presence of independent modulation in the stimulated hemisphere (**Figure 4**). This suggests that the activity at the stimulation frequency in the stimulated hemisphere consists of both a stimulation artifact and genuine independent modulation caused by 1:1 entrainment. In contrast, the non-stimulated hemisphere—used as a control since 1:1 entrainment is not expected there—exhibits only the stimulation artifact.

### 3.4 Relation between spontaneous FTG, entrained FTG and beta

#### 3.4.1 Relation between spontaneous FTG and entrained FTG

Out of 16 STN displaying sFTG, 9 STN also exhibited 1:2 eFTG. while 7 did not. In the 9 STN where sFTG and 1:2 entrainment both appeared, there was at least one contact pair per lead where they were both present.

Similarly, eFTG during stimulation was not always associated with the presence of sFTG in the stimulation OFF state, even within the same session. Indeed, in 35/44 STN with eFTG, no sFTG was recorded. Nevertheless, in specific recordings, a relation between entrained and sFTG was observed, showing a transition between spontaneous and eFTG (**Figures 6 and 7**).

**Figure 6.**
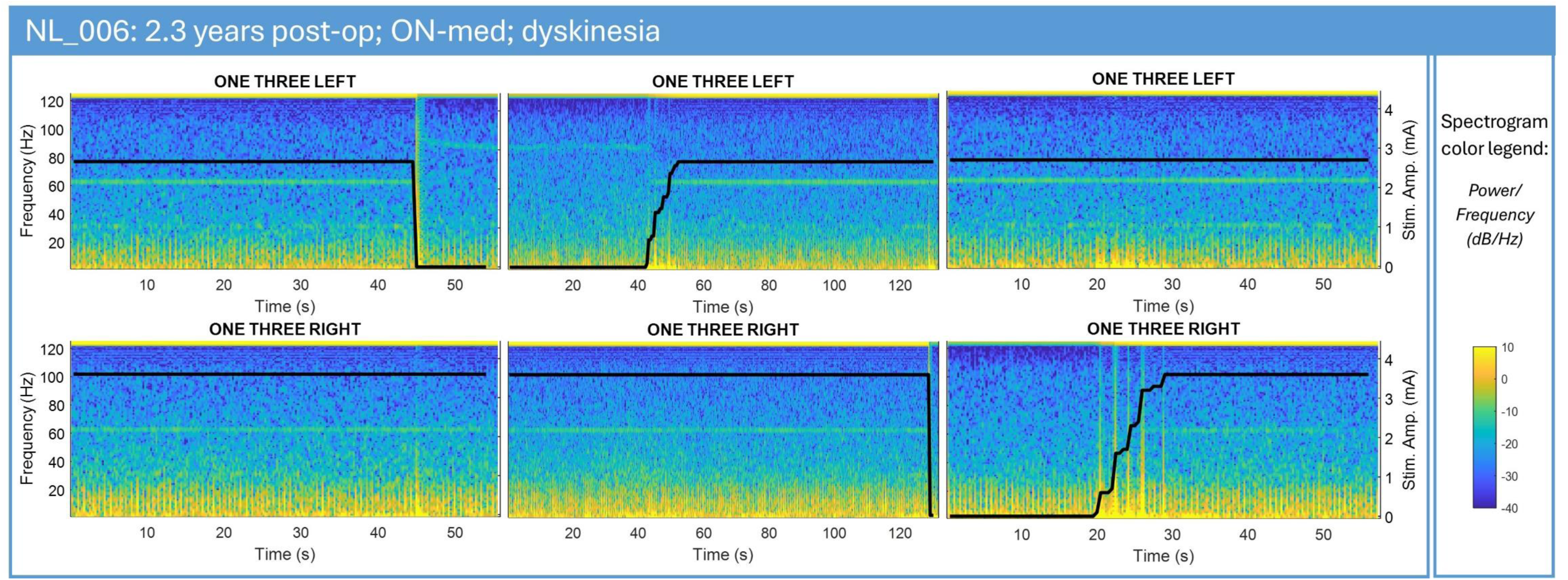
Power spectral density plots for local field potential recordings performed using BrainSense^TM^ Streaming in both hemispheres of a single patient (NL_006). Recordings were performed across three sequential sessions during a single in-clinic visit at 2 years and 4 months postoperatively, ON-medication whilst the patient was experiencing dyskinesia. In the first sessions the stimulation amplitude in the left hemisphere was decreased from 2.7mA to 0mA. During stimulation 1:2 entrained finely-tuned gamma (eFTG) was visible as a narrow band between 60-65Hz in both hemispheres. Once the stimulation was decreased in the left hemisphere the FTG frequency changed to 90Hz, suggesting a transition from 1:2 entrained to spontaneous FTG (sFTG), while remaining unchanged in the other hemisphere. In the second session the stimulation amplitude in the left hemisphere was increased from 0mA back to the original stimulation amplitude of 2.7mA. When stimulation was still set to 0mA sFTG was visible around 90Hz in the left hemisphere, which changed to narrow band activity between 60-65Hz during stimulation (1:2 eFTG), suggesting a transition from spontaneous to 1:2 eFTG. Stimulation in the right hemisphere remained constant, leading to 1:2 eFTG throughout almost the entire session. At the end of the session stimulation was decreased instantly from 3.6mA to 0mA in the right hemisphere. The 1:2 eFTG activity between 60-65Hz disappeared at this moment. In the third and final session the stimulation amplitude in the right hemisphere was slowly ramped back to the original stimulation amplitude of 3.6mA. When stimulation was set to 0mA no FTG was visible. Once stimulation was ramped up 1:2 eFTG appeared between 60-65Hz. In the left hemisphere stimulation amplitude and 1:2 eFTG were constant throughout the session. In bot hemispheres and throughout all sessions activity at the stimulation frequency showed clear relation with the stimulation amplitude and was thus considered to be of artifactual nature.

**Figure 7.**
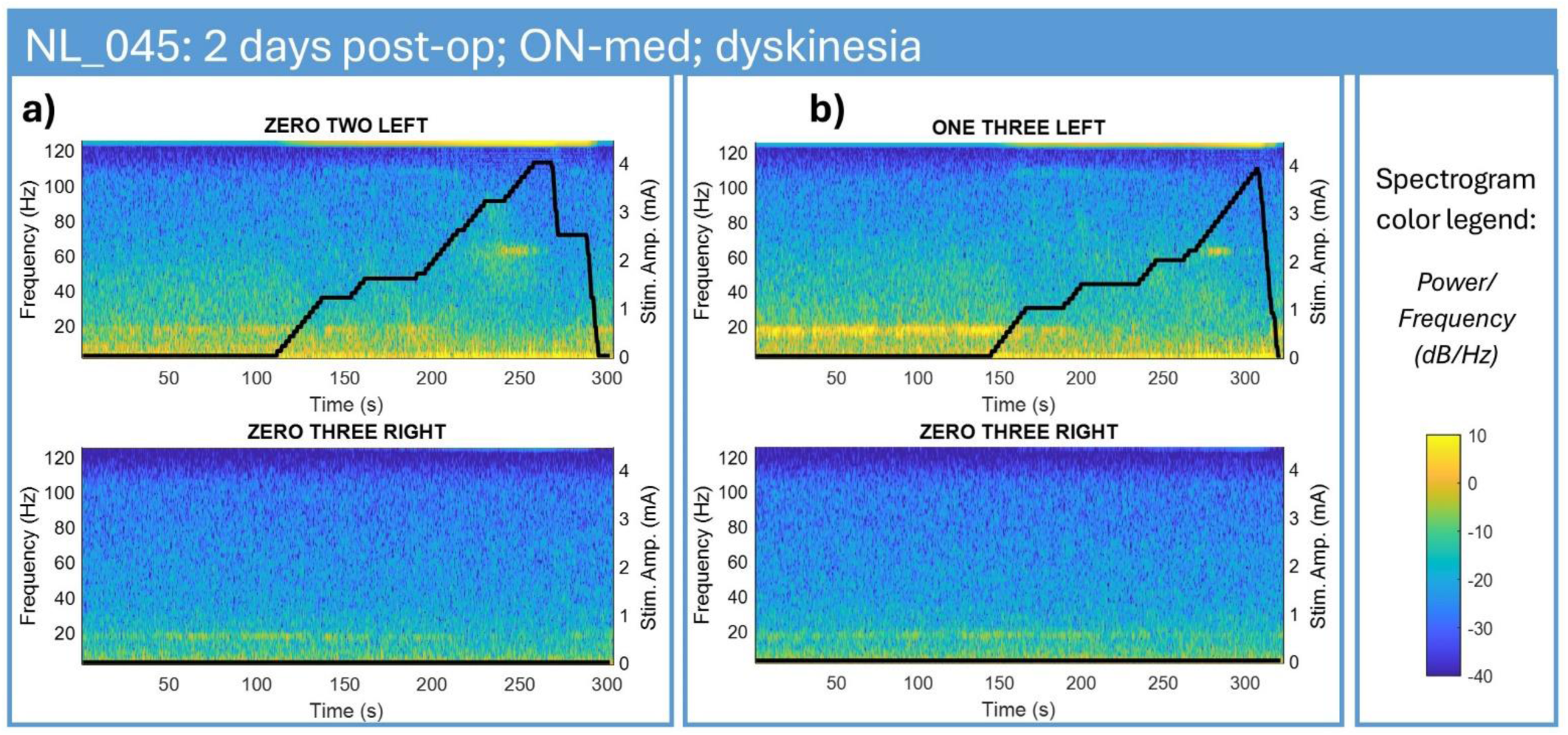
Power spectral density plots for local field potential recordings performed using BrainSense^TM^ Streaming in both hemispheres of a single patient (NL_045). Recordings were performed across two sequential sessions during a single in-clinic visit at 2 days postoperatively, ON-medication whilst the patient was experiencing dyskinesia. In the first session **(a)**, the stimulation amplitude for contact-level 1in the left hemisphere was ramped from 0mA to 4mA and back to 0mA. Before stimulation onset beta-activity was evident between 13 and 30 Hz. This beta-activity was slowly suppressed by stimulation, starting at 0.6mA, and showing complete suppression at 2.6mA. Spontaneous finely-tuned gamma (sFTG) activity occurred at a frequency-band of 104 to 110Hz during stimulation only, starting at 1.2mA, and lasting until 2.6mA stimulation. At this point sFTG transitioned to 62.5Hz, representing 1:2 entrained FTG (eFTG). When stimulation amplitude was decreased to 2.5mA 1:2 eFTG ceased, and sFTG at the 104 to 110Hz band re-occurred, however, no transitional activity could be detected. Once stimulation amplitude was reduced to 0mA sFTG ceased, and beta-activity re-occurred. Activity at the stimulation frequency showed clear relation with the stimulation amplitude and was thus considered to be of artifactual nature. Despite slight stimulation artifacts, no other influences on activity in the non-stimulated, right hemisphere, were detected. A similar pattern was seen in the second session **(b)** for stimulation through contact- level 2 in the left hemisphere.

In 11/44 STN (25%) with 1:2 entrainment, a broadband gamma activity accompanied the narrow 1:2 entrainment. This broad activity spanned a bandwidth of 15-40Hz, often showing a downward transition from 90-110Hz to 40Hz within a recording session during stimulation ramp-up (**Figure 4-A1/2 and Figure 5-C1**). This broadband gamma activity occurred in select cases only during stimulation ramping and simultaneous clinical testing (monopolar review).

#### 3.4.2 Relation between FTG and beta

In those contact pairs where spontaneous or eFTG occurred in the ON-medication state, beta-activity was present in the OFF-medication state. In 3 patients (4 STN) with sFTG, distinct beta-activity was present alongside sFTG activity in the same recordings ON-medication (**Figure 2-A)**. In all other recordings ON-medication beta-activity was low or absent when sFTG activity was present. In all STN where stimulation ramping caused 1:2 entrainment, OFF-medication, beta-suppression due to the stimulation was evident (**Figure 7**). In 11 of these 18 STN (61%) beta-suppression preceded the onset of 1:2 entrainment, whereas in 7 STN (39%) beta-suppression occurred almost simultaneously with the appearance of 1:2 entrainment.

Based on the available data, no direct relationship was found between the contact pair with the maximum beta-activity in the OFF-state and the contact pair with the maximum FTG activity in the ON-state.

## 4. Discussion

In this cohort, we characterize the sFTG activity and the clinical relations associated with 1:2 stimulation-entrained FTG activity. Additionally, we demonstrate the existence of 1:1 stimulation-entrained FTG activity at a stimulation frequency of 125Hz, describing its key features. This 1:1 entrained activity was independent of the stimulation amplitude, demonstrated an inverse relationship with 1:2 entrainment during the stimulation ON-state, and, was not observed in the non-stimulated contralateral hemisphere, all characteristics that distinguish it from a stimulation artifact.

We could not confirm the exclusive association of FTG activity with dyskinesia, since both spontaneous and eFTG were observed independently of dyskinesia and across varying clinical states. 1:2 entrainment was more frequent in patients receiving 180Hz compared to 125/130Hz stimulation. Of note, specific cases of subharmonic activity due to 180Hz stimulation were likely of artifactual origin, which warns caution in the interpretation of these findings. Beta-activity showed an inverse relation with 1:2 entrainment when transitioning between states with and without stimulation.

### 4.1 Stimulation and medication effects

Consistent with prior research, we found that sFTG predominantly appeared in the ON-medication state [30], although our findings also demonstrate that both spontaneous and eFTG can emerge OFF-medication, supporting earlier findings that FTG may spontaneously align with effective DBS frequencies regardless of medication status [31, 32]. In this population, sFTG was found to occur between 63 and 110Hz, mostly in the OFF stimulation state. However, it occasionally also emerged during (low-amplitude) stimulation, before entrained 1:2 activity occurred, suggesting a potential correlation with the therapeutic threshold (ON state), independent of the effective treatment (i.e., stimulation or medication).

In previous studies, FTG has been largely linked to dyskinesia [18, 21, 33] and has even been referred to as "harmful entrainment" [34]. However, our findings support and expand the evidence that both spontaneous and eFTG can occur independently of dyskinesia [23, 26, 30, 32], highlighting the need to reassess FTG’s clinical relevance beyond manifest motor symptoms. No consistent link was found between patient, disease, or DBS characteristics and (entrained) FTG, except for its strong association with stimulation and medication states.

Still, some patients did not show eFTG even under high stimulation or ON-medication, and despite having dyskinesia, suggesting other potential contributing factors. Previous studies suggest that STN lead location, individual surgical recovery (e.g., stun effect), and disease progression may play a role [35, 36]. Our data, however, did not always support this. Therefore, these hypotheses merit more research.

We also found that higher-frequency stimulation (180 vs. 125Hz) led more frequently to 1:2 entrainment (55% vs. 28% of STN), independent of tremor, which potentially reflects the effect of increased stimulation energy, with a less selective neuronal engagement, or the physiologically higher firing rates of STN neurons[37]. In addition, considering that all the patients receiving 180Hz stimulation had tremor, and those receiving 125Hz did not, this could reflect a selective response of tremor-related neurons which may be more prone to entrain with high stimulation frequencies. These observations are supported by clinical practice, where higher stimulation frequencies tend to lead to side-effects at lower stimulation amplitudes and are more effective in suppressing tremor. Although some hypotheses suggest that high-frequency stimulation (e.g. ≥ 130Hz) may reduce the chance of 1:2 entrainment, due to axonal or synaptic failure [25, 38], our data, and data from preclinical studies [39], show that high stimulation frequencies require lower stimulation amplitude to achieve similar stimulation effects compared to lower frequency stimulation. This finding aligns with the principles of the ‘Arnold’s tongue’ framework [25], which predicts that 180Hz stimulation can induce 1:2 entrainment at lower stimulation amplitudes. In some cases, subharmonic 90Hz activity during 180Hz stimulation was also present at 0mA and was not modulated by stimulation amplitude. In these instances, the subharmonic activity was likely artifactual. This may arise because the Percept^TM^ PC system is optimized for recording physiological signals free from stimulation artifacts within a frequency range up to 80Hz [29]. Since 90Hz falls outside this range, the system may have limitations in recording at this frequency, making it more susceptible to stimulation artifacts.

Our findings reveal that both sFTG and eFTG are inherently unstable, fluctuating even during constant medication or stimulation settings. In several cases, 1:2 entrainment ended before stimulation amplitude was lowered or even during ramping up of the stimulation, suggesting that desynchronization at half the stimulation frequency may result from increased amplitudes or synaptic fatigue from high-frequency stimulation [25, 38] or may be modulated by other factors such as movement, speech, or attention.

### 4.2 Inter-frequency relations

The activity around the stimulation frequency is generally considered a stimulation artifact. However, here we argue that this activity is not always solely of artifactual nature. We found that activity at the stimulation frequency in the stimulated hemisphere can consists of both a stimulation artifact as well as a modulation independent from the stimulation amplitude, which we consider as 1:1 entrainment. In line with this hypothesis, the non-stimulated hemisphere where 1:1 entrainment is not expected, exhibits only the stimulation artifact and no independent modulation.

Additionally, the independent modulations which we considered 1:1 entrainment showed an inverse relation to 1:2 entrainment activity, suggesting a competitive interaction between these rhythms, possibly due to shared neural pathways. In most cases, 1:2 entrainment cessation corresponded to increased 1:1 entrainment amplitudes whereas stronger 1:2 entrainment was often linked to reduced 1:1 entrainment power. This aligns with the 1:1 entrainment model proposed by Sermon et al. (2023) [25], and is supported by cellular imaging studies showing strong membrane depolarizations at stimulation frequencies (1:1 entrainment) in rodents, in recordings made free from stimulation artifacts [39]. The presence of 1:1 entrainment for 180Hz stimulation could not be evaluated here because of the sampling rate of 250hz inherent to the Percept^TM^ PC system. Taken together, these observations reinforce the physiological basis of eFTG and support previous findings that DBS-induced entrainment depends on a fine balance between stimulation parameters and neuronal dynamics [30, 32].

In a subset of the patients with bilateral recordings during 125Hz stimulation ramping from 0–4mA in one hemisphere, activity at the stimulation frequency was detected in the non-stimulated STN. Because this activity mirrored changes in stimulation amplitude in the stimulated side, this suggests rather an artifact than interhemispheric effects of stimulation, which have been previously shown [40, 41]. Indeed, no other physiological effects, such as beta-suppression or 1:2/1:1 entrainment, were observed in the non-stimulated STN. It remains unclear whether this cross-hemispheric stimulation activity caused symptom relief. Importantly, when assessing 1:1 entrainment, artifacts due to contralateral stimulation must be considered to avoid misinterpretation of artifacts as genuine physiological responses.

In several cases of 1:2 entrainment, we also observed broadband gamma activity, with a bandwidth of 15–40Hz, shifting from higher (∼100Hz) to lower (∼40Hz) frequencies within single sessions. Similar broadband activity has been linked to contralateral movements [42, 43], suggesting that in our recordings this activity might be caused by patients performing motor tasks (e.g., finger tapping) during monopolar reviews [26]. Alternatively, the broadband activity may reflect a transition between spontaneous and eFTG, as proposed by Mathiopoulou et al. (2025) [26], however, this hypothesis does not explain persistent activity during 1:2 entrainment or at frequencies below the 1:2 entrainment frequency, as shown here. Our data also confirmed a consistent inverse relation between beta-activity and FTG, particularly eFTG, supporting an antagonistic relationship [24, 32]. Interestingly, sFTG co-occurred with beta peaks in 3 patients (4 STNs), nonetheless, beta-suppression may still have occurred. Future work should explore the mechanisms linking beta and FTG.

The dynamic interplay between beta and FTG suggests eFTG could be a useful physiomarker for aDBS, complementing beta-activity for more tailored therapy. Unlike beta or sFTG, eFTG’s characteristics of a constant and predictable frequency render it a particularly promising physiomarker. A combined beta–FTG approach may improve aDBS algorithms, offering more precise symptom control across diverse patients [17, 21, 23, 26, 32]. However, as not all patients exhibit eFTG, personalized strategies remain essential.

### 4.3 Limitations and future directions

One limitation of this study is its retrospective design, which did not include standardized experimental protocols or extension of the recordings to further explore some of the observed phenomena. On the other hand, using recordings as they are collected in a naturalistic clinical setting has allowed us to study conditions that might have been missed in a predefined study protocol (such as stimulation at 180Hz or the neurophysiological consequences of stun effect). These preliminary observations should be expanded in future structured, prospective protocols to more extensively assess and confirm the different (entrained) FTG characteristics shown here across different conditions, as well as the relations with disease characteristics.

### 4.4 Conclusions

This study provides novel insights into the prevalence, dynamics, and clinical significance of FTG in patients with PD undergoing STN-DBS. Adding to the most recent observations, our findings demonstrate that FTG—both spontaneous and entrained—can occur independently of dyskinesia and during varying clinical states.

Importantly, here we demonstrate the possibility of recording 1:1 entrainment and confirm its interaction with 1:2 entrainment in line with the ‘Arnold’s tongue’ framework. We also emphasize the inherent instability of spontaneous and eFTG and its complex interactions with other oscillatory activities such as beta rhythms.

Crucially, understanding the interactions between beta-activity, sFTG, and 1:2 and 1:1 entrainment, and their relation with PD symptoms, could advance the use of combined physiomarkers to enable more effective and personalized aDBS protocols. However, as highlighted in this study, the observed selective occurrence of eFTG in some patients, the possible presence of artifacts overlapping with the neurophysiological features, and the cross-hemisphere influences reinforce the need for a cautious and tailored approach.

## Data availability

The data that support the findings of this study are available from the corresponding author upon reasonable request.

## Data Availability

All data produced in the present study are available upon reasonable request to the authors

## Acknowledgments

The authors are thankful to Dr. E. Lowet, G. Leogrande, and Prof. Dr. J.J. van Hilten for their helpful insights.

## Authors’ Roles

- Marjolein Muller: design, execution, writing, and editing of the final version of the manuscript
- Saskia van der Gaag: execution, and editing of the final version of the manuscript
- Rodi Zutt: execution, and editing of the final version of the manuscript
- Eline Rouleau: editing of the final version of the manuscript
- Carel Hoffmann: editing of the final version of the manuscript
- Niels van der Gaag: editing of the final version of the manuscript
- Thomas van Essen: editing of the final version of the manuscript
- Alfred Schouten: design, and editing of the final version of the manuscript
- Fiorella Contarino: design, execution, writing, and editing of the final version of the manuscript

